# Variant Subpopulations in Nitroimidazole Resistance Genes in DELIBERATE trial participants

**DOI:** 10.1101/2025.09.15.25335512

**Authors:** Fahd Naufal, Megan L Folkerts, Jason D Limberis, Elizabeth M Streicher, Alina Nalyvayko, Gary Maartens, Kelly E. Dooley, Robin M. Warren, David Engelthaler, John Z. Metcalfe

## Abstract

Targeted next-generation sequencing (tNGS) enables rapid detection of drug resistance in Mycobacterium tuberculosis (Mtb), including minor variants that may indicate emerging resistance. We applied tNGS to isolates from 34 (of 49 eligible) participants in a randomized trial of bedaquiline (BDQ) and delamanid (DLM) co-administration for multidrug-resistant TB (MDR-TB). We detected baseline nonsynonymous variants in the nitroimidazole resistance genes *fgd2* and *ddn* (BDQ arm) and *fbiB* (DLM arm) among 2/12 and 2/12 patients, respectively, and emergent subpopulations with nonsynonymous variants in *fbiB, fbiC*, and *ddn* as early as week 1 in 3/34 patients. These findings demonstrate that variants in nitroimidazole resistance genes may be present in the absence of drug exposure, and that tNGS can effectively track variant emergence during MDR-TB treatment.

---

Targeted next-generation sequencing (tNGS) for rapid drug susceptibility testing (DST) is an emerging approach to address the global *Mycobacterium tuberculosis* (Mtb) burden through enabling the identification of both known and novel mutations linked to drug resistance. This allows for timely predictions of drug susceptibility, supporting the design of tailored treatment regimens to improve patient outcomes. Additionally, tNGS’s capability to detect minor variants may elucidate the early emergence of acquired resistance.

Heteroresistance in multi drug-resistant TB (MDR-TB) complicates both diagnostic and treatment strategies.^1-3^ It describes the presence of Mtb subpopulations that display varying degrees of drug susceptibility, even while conventional phenotypic testing classifies an infection as uniformly susceptible or resistant. In this study, we leveraged a randomized trial assessing bedaquiline-delamanid co-administration (ACTG A5343; NCT02583048) to perform tNGS on Mtb isolates from MGIT (mycobacterial growth indicator tube) cultures to assess longitudinal emergence of genotypically variant subpopulations within bedaquiline and nitroimidazole resistance genes. We used a previously described tNGS pipeline^4^ to detect variants in *mmpR5*. Genes known to be involved in nitroimidazole resistance (*ddn, fgd1, fgd2, fbiA, fbiB*, and *fbiC*) were sequenced using a tiled amplicon approach to cover each target, as previously described.^5, 6^ Briefly, primer sequences (Supplemental Table 1) included a universal tail sequence, which facilitated the addition of an Illumina adapter in a second polymerase chain reaction step. Quality controlled libraries were pooled equimolarly and sequenced on an Illumina NextSeq1000 with 300 bp, paired-end chemistry. All sequencing runs included at least 30% phiX sequencing control (Illumina) to facilitate base diversity. Data analysis was performed using the TB Amplicon Sequencing Analysis Pipeline (ASAP) with Single Molecule Overlapping Read technology (SMOR).^5, 6^ Briefly, the ASAP-SMOR pipeline requires that overlapping paired reads agree for a variant call to be made. By doing so, sequencing error is reduced, increasing the confidence of variant calls. The pipeline requires a minimum of five paired reads to make an individual variant call. For this dataset, variant calls were made down to 1%; therefore, the minimum depth needed for variant calling was 500 paired reads. Variants were analyzed using R (4.3.0) and the packages “ggpubr” (0.6.0), “Hmisc” (5.1-0), and “tidyverse” (2.0.0).

Forty-nine participants from the A5343 trial with MDR-TB were assessed for eligibility, of whom three participants had no isolates that could not be subcultured and were not sequenced. Of the remaining 46 participants, a further 10 had only heat-inactivated lysates available for sequencing and were excluded due to the presence of numerous ultra-small variant calls not seen in isolates with higher quality DNA extraction; 2/46 (4%) isolates did demonstrate contamination with non-tuberculous mycobacteria. Participants received either bedaquiline (BDQ) only (*n*=12), the nitroimidazole delamanid (DLM) only (*n*=12), or BDQ plus DLM (*n*=10), in addition to a multi-drug background therapy (MBT) for a median of 12 days prior to initiation of study medications. MBT comprised a total of 4-5 additional agents, excluding clofazimine and moxifloxacin due to their effects on QT prolongation. Patient treatment adherence was assured as the A5343 participants had a period of inpatient management and trial directly observed therapy during study drug administration.

Within *mmpR5*, only the c.-11C>A promoter mutation was detected among 16/34 (47%) A5343 participants; among 2 (13%) of whom the mutation was not detected after the week 1 visit. Among genes associated with nitroimidazole resistance, after excluding lineage markers (L2.2.1, fgd1_c.960T>C; L4.1.2.1, fgd1_c.809A>T), we identified fifteen total mutations across all time points, seven of which were synonymous (**Figure 1**).^7, 8^ One synonymous mutation in fbiB_c.273C>G was previously reported in the WHO Catalog and graded as uncertain significance.^8^ Two participants in both the BDQ arm (16.7%; *n*=2/12) and the DLM arm (16.7%; *n*=2/12) had fixed mutations with unknown association with resistance at baseline. Where longitudinal samples were available, an additional two participants (16.7%; *n*=2/12) in the DLM arm acquired genotypically variant subpopulations with mutations in *fbiB* and *fbiC* genes over time (Table 1); one of these (PID#20) occurred at week 24. In the BDQ plus DLM arm, no participants had baseline mutations, though subpopulations with mutations in *ddn* gene were demonstrated for one participant at week 4; no further culture-positive timepoints were available for this participant.

**Figure 1.**
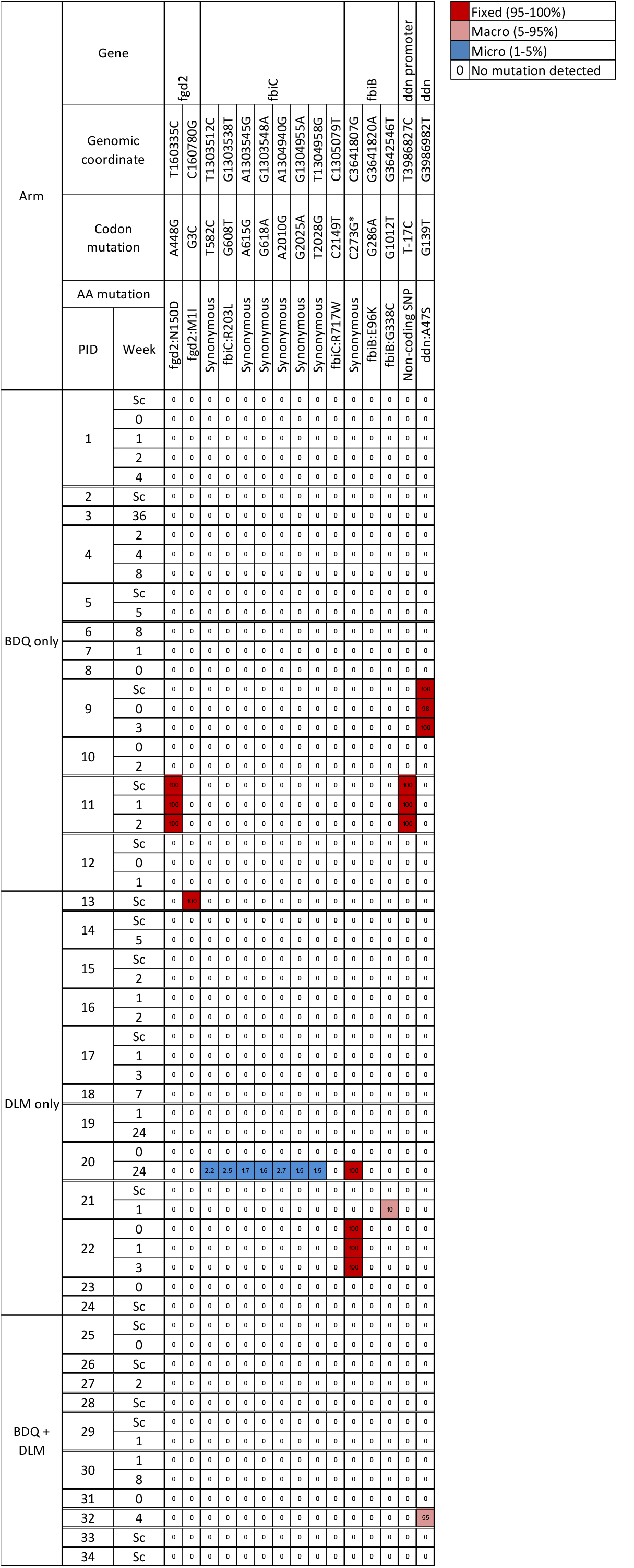
Genotypic variant subpopulations in nitroimidazole resistance genes. Participants from the A5343 trial with multidrug-resistant TB were randomized to receive BDQ only, DLM only, or BDQ plus DLM, in addition to a multidrug background therapy. No mutations in *Rv0678* associated with bedaquiline resistance were noted. Two previously described lineage markers (L2.2.1, *fgd1*_c.960T>C; L4.1.2.1, *fgd1*_c.809A>T) were removed. The emergence of genotypic variants in two participants (*fbiC*_c.608G>T in PID#20 and *fbiB*_c.1012G>T in PID#21) occurred in the DLM only arm. In the BDQ only arm, fixed mutations with unknown association with resistance were identified in one participant in *ddn* (*ddn*_c.139G>T), and in another participant in *fgd2* (*fgd2*_c.448A>G) and the *ddn* promoter region (c.-17T>C). One participant in the DLM only arm had a fixed mutation with unknown association with resistance in *fbiB* (*fbiB*_c.273C>G). One participant in the BDQ plus DLM arm had genotypically variant subpopulations in *ddn* (*ddn*_c.139G>T). Sc – Screening visit; AA – Amino acid; BDQ – Bedaquiline; DLM – Delamanid; a 0 in the heatmap indicates adequate coverage at the position, but no SNP.

Heteroresistance in Mtb can arise from selective pressures imposed by inconsistent or incomplete treatment regimens or the intrinsic genetic diversity of transmitted or pre-existing mycobacterial populations. In this study, genotypically variant subpopulations with mutations in *fbiB* and *fbiC* genes emerged at week 1 (PID#21) and week 24 (PID#20) of treatment, respectively.^9^ Baseline “fixed” mutations were observed in *fgd* and *ddn* in participants in the BDQ only arm, and in *fbiB* in the DLM only arm. Further investigation with a combination of genotypic, phenotypic, and structural analysis will yield more information on the roles of these specific mutations.^10^

We demonstrate a tNGS approach able to detect and quantify variant subpopulations of mycobacteria down to 1% of the total population. Among patients with both MDR-TB enrolled in a clinical trial in South Africa and Peru, with no documented exposure to nitroimidazoles, we found populations of Mtb with fixed nonsynonymous variants in the *fgd2* and *ddn* genes at baseline. Further, in patients with MDR-TB, we describe at least two instances of emergence of nonsynonymous variants in *fbiB* and *fbiC*. A major limitation of our report is that phenotypic drug susceptibility testing for delamanid was not performed, and therefore the association with drug resistance is unknown. In addition, the availability of sequential viable isolates was inconsistent, and therefore consecutive sampling timepoints were often unavailable, and potentially missing not at random. Nevertheless, given the importance of nitroimidazoles to current and future TB regimens, our report highlights the possibility of a basal rate of both pre-existing variants in genes associated with nitroimidazole resistance, as well as the emergence of such variants even under controlled trial conditions.

## Data Availability

All data produced in the present study are available upon reasonable request to the authors

## Notes

### Competing Interest Statement

The authors have declared no competing interest.

### Clinical Trial

NCT02583048

### Funding Statement

This study was funded by the National Institutes of Health (R01AI136894)

### Author Declarations

Ethics committee of the University of California San Francisco and the Stellenbosch University Health Research Ethics Committee gave ethical approval for this work

### Summary of Updates

We have corrected a spelling error in the author names.

## References

1. Chen Y, Ji L, Liu Q, Li J, Hong C, Jiang Q, et al. Lesion Heterogeneity and Long-Term Heteroresistance in Multidrug-Resistant Tuberculosis. J Infect Dis. 2021;224(5):889–93.

2. Liu Q, Via LE, Luo T, Liang L, Liu X, Wu S, et al. Within patient microevolution of Mycobacterium tuberculosis correlates with heterogeneous responses to treatment. Sci Rep. 2015;5:17507.

3. Bernard C, Aubry A, Chauffour A, Brossier F, Robert J, Veziris N. In vivo Mycobacterium tuberculosis fluoroquinolone resistance emergence: A complex phenomenon poorly detected by current diagnostic tests. J Antimicrob Chemother. 2016;71(12):3465–72.

4. Derendinger B DA, de Vos M, Huo S, Alberts R, Tadokera R, Limberis J, Sirgel F, Dolby T, Spies C, Reuter A, Folkerts M, Allender C, Lemmer D, Van Rie A, Gagneux S, Rigouts L, Te Riele J, Dheda K, Engelthaler DM, Warren R, Metcalfe J, Cox H, Theron G. Bedaquiline resistance in patients with drug-resistant tuberculosis in Cape Town, South Africa: a retrospective longitudinal cohort study. Lancet Microbe. 2023;4(12):e972–e82.

5. Colman RE, Anderson J, Lemmer D, Lehmkuhl E, Georghiou SB, Heaton H, et al. Rapid Drug Susceptibility Testing of Drug-Resistant Mycobacterium tuberculosis Isolates Directly from Clinical Samples by Use of Amplicon Sequencing: a Proof-of-Concept Study. J Clin Microbiol. 2016;54(8):2058–67.

6. Colman RE, Schupp JM, Hicks ND, Smith DE, Buchhagen JL, Valafar F, et al. Detection of Low-Level Mixed-Population Drug Resistance in Mycobacterium tuberculosis Using High Fidelity Amplicon Sequencing. PLoS One. 2015;10(5): e0126626.

7. Reichmuth ML, Hömke R, Zürcher K, Sander P, Avihingsanon A, Collantes J, et al. Natural Polymorphisms in Mycobacterium tuberculosis Conferring Resistance to Delamanid in Drug-Naive Patients. Antimicrob Agents Chemother. 2020;64(11):e00513–20.

8. World Health Organization. Catalogue of Mutations in Mycobacterium Tuberculosis Complex and Their Association with Drug Resistance, 2nd ed.: World Health Organization; 2023 [Available from: https://www.who.int/publications/i/item/9789240082410.

9. Villellas C, Coeck N, Meehan CJ, Lounis N, De Jong B, Rigouts L, et al. Unexpected high prevalence of resistance-associated Rv0678 variants in MDR-TB patients without documented prior use of clofazimine or bedaquiline. Journal of Antimicrobial Chemotherapy. 2017;72(3):684–90.

10. Battaglia S, Spitaleri A, Cabibbe AM, Meehan CJ, Utpatel C, Ismail N, et al. Characterization of genomic variants associated with resistance to bedaquiline and delamanid in naive mycobacterium tuberculosis clinical strains. J Clin Microbiol. 2020;58(11):e01304–20.

